# Mental State of Inpatients with COVID-19: a Computational Psychiatry Approach

**DOI:** 10.1101/2021.08.15.21262025

**Authors:** M.Yu. Sorokin, E.I. Palchikova, A.A. Kibitov, E.D. Kasyanov, M.A. Khobeysh, E.Yu. Zubova

## Abstract

The overload of healthcare systems around the world and the danger of infection have limited the ability of researchers to obtain sufficient and reliable data on psychopathology in hospitalized patients with COVID-19. The relationship between severe SARS-CoV-2 infection and specific mental disturbances remains poorly understood.

**Aim:** to reveal the possibility of identifying the typology and frequency of psychiatric syndromes associated with acute COVID-19 using cluster analysis of discrete psychopathological phenomena.

**Materials and methods:** Descriptive data on the mental state of 55 inpatients with COVID-19 were obtained by young-career physicians with psychiatric backgrounds. Classification of observed clinical phenomena was performed with k-means cluster analysis of variables codded from the main psychopathological symptoms. Dispersion analysis with p-level 0.05 was used to reveal the cluster’s differences in demography, parameters of inflammation and respiration function collected on the basis of the original medical records.

**Results:** Three resulting clusters of patients were identified: persons with anxiety, disorders of fluency and tempo of thinking, mood, attention, motor-volitional sphere, reduced insight, and pessimistic plans for the future (n=11); persons without psychopathology (n=37); persons with disorientation, disorders of memory, attention, fluency, and tempo of thinking, reduced insight (n=7). The development of a certain type of impaired mental state was specifically associated with: age, lung lesions according to computed tomography, saturation, respiratory rate, C-reactive protein level, platelet count.

**Conclusion:** The prevalence and typology of psychiatric disorders in patients with acute COVID-19 were described using the computational psychiatry approach.

## INTRODUCTION

Neurotropic nature of SARS-CoV-2 predetermines psychiatric disorders in some patients with COVID-19 [1-3]. However, most publications on the psychological and mental impact of COVID-19 present the results of online and cross-sectional studies of the general population [4-7] or post-recovery data of patients who have suffered the acuteSARS-CoV-2 infection earlier [8-11]. Even the clinical findings from previous coronavirus crises are mostly symptom- and dimension-oriented. [12, 13].

The complex clinical picture and frequency of psychiatric syndromes in patients with current SARS-CoV-2 infection remain poorly understood. [14, 15]. A few studies present case reports of rare psychiatric conditions [16-18]. Some data were published about the existence of neurological disturbances in hospitalized patients with COVID-19 [19-21]. Few studies are systematic assessments of the mental status of inpatients with COVID-19 [22]. All these results are often obtained by non-psychiatric health professionals. At the same time, neuropsychiatric disorders are a COVID-19 death risk factor [22, 23], so they need to be diagnosed in a timely manner and appropriately treated. In this case, the lack of data on typical mental status variations in COVID-19 patients must be addressed because of the importance of this phenomenological information as a potential target for clinical screening and risk assessment by general practitioners.

At the same time, the extreme overload of healthcare systems around the world and the danger of infection have limited the ability of psychiatric researchers to obtain sufficient and reliable data on psychopathology in hospitalized patients with COVID-19. The relationship between severe infection and specific psychiatric syndromes remains to be explored. Back in the early days of psychiatry as a medical specialty, solving similar problems associated with syphilis and progressive paralysis took more than 100 years [24]. Computational psychiatry is considered a promising methodology for assessing complex clinical events with a large number of factors and predictors that can lead to ambiguous clinical conditions in patients [25,26]. An important aspect of this approach is verification of the observed mental disturbances using certain pathogenetic indicators, such as inflammation and abnormalities of physiological functions [27].

The hypothesis of the study: nervous system damage caused by SARS-Cov-2 can have a variety of psychopathological manifestations in patients and must be associated with specific clinical parameters.

Aim of the study: to reveal the possibility of identifying the typology and frequency of psychiatric syndromes associated with acute COVID-19 using cluster analysis of discrete psychopathological phenomena.

## MATERIALS AND METHODS

Assessment of the mental state of patients with COVID-19 requires specialized education and sufficient clinical practice of a physician. These requirements are unattainable in the real world of the COVID-19 crisis. We have obtained descriptive data on the mental state of 55 inpatients with COVID-19 captured by trainees of the V.M. Bekhterev NMRC for Psychiatry and Neurology, Saint-Petersburg, Russia. Young career physicians with psychiatric backgrounds should provide enough quality in the process of data acquisition. To standardize mental state assessment and to maximize inter-rater reliability, discrete psychopathological phenomena were preidentified for raters. They used a scale from 0 to 1 points, where 0 - absence and 1 - presence of violations. The possible range of severity between 0 and 1 point should provide “artifact correction” during data acquisition, and k-means cluster analysis of quantitative variables codded from the main psychopathological symptoms allowed to perform classification of observed clinical phenomena. It is considered that quality control during data acquisition, artifact correction, and robust statistical algorithms are essential for computational technologies in psychiatry [28]. The t-Student’s and the U-Mann-Whitney tests with p-level 0.05 were used to reveal cluster’s differences in parameters of inflammation and respiration function which were suggested as a physiological background of psychopathology in COVID-19 patients. Clinical parameters for the patients were collected on the basis of the original medical records. Descriptions of subgroups were presented in means M[S.D.] or medians Me(IQR).

The study design was controlled by the independent ethical committee of the V.M. Bekhterev National Medical Research Center for Psychiatry and Neurology, Russian Federation Health Ministry. It was in conformity with the Helsinki Declaration and the standard of good clinical practice (GCP). It included collection of anamnestic, socio-demographic data, clinical parameters based on the original medical records after the patients signed a voluntary informed consent and their current mental state was tested.

Inclusion criteria: 1) ability to read, understand and readiness to sign a voluntary informed consent to take part in the study; 2) a hospitalization due to COVID-19 diagnosis; 3) ability to fulfil the study procedures.

Non-inclusion criteria: 1) extremely high severity of the current condition with insufficient respiratory function; 2) Age less than 18 years. Exclusion criteria: refusal to comply with the study procedures at any stage of the study.

## RESULTS

The sample of the patients consisted of 21 men and 34 women, with a mean age of 51.5[20.9] years. Higher and not completed higher education was characteristic of 30 patients (54.5%), secondary education – of 11 patients (20.0%), primary education – of 14 patients (25.5%). The majority of the sample were married people – 33 (60%), and the smaller share were single persons – 21 patients (38.2%). Also, the majority of the patients studied or worked full time – 31 (56.4%), and the smaller share were unemployed – 23 (41.8%). Data about education and occupation for 1 patient (1.8%) were missing. The most prevalent comorbidities were cardiovascular disorders – 11 patients (20.0%), then endocrinological disorders – 6 (10.6%), gastrointestinal – 5 (9.1%), respiratory – 2 (3.6%), renal and neurological disorders were the rarest – in 1 patient (1.8%) for each comorbidity. The mean percentage of lung lesions according to computed tomography data was 20.1%[19.1] and saturation lower than 95% was characteristic of 16 patients.

Three resulting clusters of patients were identified (without differences in gender, somatic and mental comorbidities). 1st (n=11): patients with anxiety, disorders of fluency and tempo of thinking, mood, attention, motor-volitional sphere, reduced insight, and pessimistic plans for the future. 2nd (n=37): patients without psychopathology. 3d (n=7): patients with disorientation, disorders of memory, attention, fluency, and tempo of thinking, reduced insight.

Representatives of cluster 1, in comparison with cluster 2 (without mental disturbances, had more lung lesions according to computed tomography: 20%(34) vs. 15%(18), p=0.018. There were no significant differences in saturation, respiratory rate, and other laboratory parameters, as well as in age between patients from cluster 1 and cluster 2.

Other patients with mental abnormalities (cluster 3) were older: 76.9[14.7] versus healthy patients (cluster 2) 50.9[17.8], p=0.001, as well as versus patients with anxiety and mood disturbances (cluster 1) 60.9[24.3], p=0.027. Cluster 3 patients, in comparison with cluster 2 (patients without mental abnormalities), were clinically different by a more severe course of the disease based on the results of laboratory and instrumental methods: a higher percentage of lung damage (31%(35) vs. 15%(18), p<0.001); higher level of C-reactive protein (126mg/L(236) vs. 10mg/L(21), p<0.001); lower saturation (89%(13) vs. 97%(4), p<0.001), higher respiratory rate (21(6) vs. 18(4), p<0.001).

Patients from cluster 3 versus cluster 1 clinically differed: a higher percentage of lung lesions on computed tomography (31%(35) vs. 25%(34), p=0.029); higher C-reactive protein level (126mg/L(236) vs. 16mg/L(88), p<0.001); lower saturation (89%(13) vs. 95.5%(4), p=0.005); higher respiratory rate (21/min.(6) vs. 19/min.(7), p=0.035); lower platelet count (139*109/L(129) vs. 322*109/L(129), p=0.006).

## DISCUSSION

To our knowledge, this is the first study performed using the computational psychiatry approach to assess the presence and typology of psychiatric disorders in patients with acute COVID-19. The hypothesis of the study was confirmed: differences in the presence of psychopathology and the development of a certain type of impaired mental state were associated with specific clinical and laboratory parameters of patients. The combined representation of anxiety and depressive symptoms with psychomotor retardation were characteristic of 20% of inpatients with acute COVID-19. Symptoms of impaired consciousness and memory, combined with impaired insight, were present in 13% of the sample.

The study had several limitations. Firstly, patients in extremely severe current condition with insufficient respiratory function were not included in the study, although they could have more pronounced mental disturbances. Second limitation was the small size of the sample due to the limited access to COVID-19 patients. Thirdly, standardized psychiatric diagnostic methods and tests were not used because of the lack of time and acute infection process in the study participants. Lastly, assessment of the mental state of patients with COVID-19 was conducted by psychiatric trainees without psychiatric medical licenses yet.

The results of the study should be used for better risk assessment of people with coronavirus infection and prediction of neuropsychiatric consequences as a marker of more unfavorable course of the disease.

## Data Availability

The data that support the findings of this study are available from the corresponding author upon reasonable request.

## References

1. van Vuren, E. J., Steyn, S. F., Brink, C. B., Möller, M., Viljoen, F. P., & Harvey, B. H. (2021). The neuropsychiatric manifestations of COVID-19: Interactions with psychiatric illness and pharmacological treatment. Biomedicine & Pharmacotherapy, 111200. doi: 10.1016/j.biopha.2020.111200

2. Afshar-Oromieh, A., Prosch, H., Schaefer-Prokop, C., Bohn, K. P., Alberts, I., Mingels, C., … & Ebner, L. (2021). A comprehensive review of imaging findings in COVID-19-status in early 2021. European journal of nuclear medicine and molecular imaging, 1–25. doi: 10.1007/s00259-021-05375-3

3. Khosroshahi, L. M., Rokni, M., Mokhtari, T., & Noorbakhsh, F. (2021). Immunology, immunopathogenesis and immunotherapeutics of COVID-19; an overview. International Immunopharmacology, 93, 107364. doi: 10.1016/j.intimp.2020.107364.

4. Deng, J., Zhou, F., Hou, W., Silver, Z., Wong, C. Y., Chang, O., … & Zuo, Q. K. (2020). The prevalence of depression, anxiety, and sleep disturbances in COVID□19 patients: a meta□analysis. Annals of the New York Academy of Sciences. doi: 10.1111/nyas.14506

5. Krishnamoorthy, Y., Nagarajan, R., Saya, G. K., & Menon, V. (2020). Prevalence of psychological morbidities among general population, healthcare workers and COVID-19 patients amidst the COVID-19 pandemic: A systematic review and meta-analysis. Psychiatry research, 293, 113382. doi: 10.1016/j.psychres.2020.113382

6. Cuiyan, W., Riyu, P., Xiaoyang, W., Yilin, T., & Linkang, X. (2020). McIntyre Roger S, Choo Faith N, Tran Bach, Ho Roger, Sharma Vijay K, Ho Cyrus. A longitudinal study on the mental health of general population during the COVID-19 epidemic in China. Brain Behav Immun, 87, 40–48. doi: 10.1016/j.bbi.2020.04.028

7. Jones, E. A., Mitra, A. K., & Bhuiyan, A. R. (2021). Impact of COVID-19 on mental health in adolescents: a systematic review. International journal of environmental research and public health, 18(5), 2470. doi: 10.3390/ijerph18052470

8. Varatharaj, A., Thomas, N., Ellul, M. A., Davies, N. W., Pollak, T. A., Tenorio, E. L., … & Plant, G. (2020). Neurological and neuropsychiatric complications of COVID-19 in 153 patients: a UK-wide surveillance study. The Lancet Psychiatry, 7(10), 875–882. doi: 10.1016/S2215-0366(20)30287-X

9. Rogers, J. P., & David, A. S. (2021). A longer look at COVID-19 and neuropsychiatric outcomes. The Lancet Psychiatry, 8(5), 351–352. doi: 10.1016/S2215-0366(21)00120-6

10. Hampshire, A., Trender, W., Chamberlain, S. R., Jolly, A. E., Grant, J. E., Patrick, F., … & Mehta, M. A. (2021). Cognitive deficits in people who have recovered from COVID-19. EClinicalMedicine, 101044. doi: 10.1016/j.eclinm.2021.101044

11. Bennett, K. E., Mullooly, M., O’Loughlin, M., Fitzgerald, M., O’Donnell, J., O’Connor, L., … & Cuddihy, J. (2021). Underlying conditions and risk of hospitalisation, ICU admission and mortality among those with COVID-19 in Ireland: A national surveillance study. The Lancet Regional Health-Europe, 5, 100097. doi: 10.1016/j.lanepe.2021.100097

12. Rogers, J. P., Chesney, E., Oliver, D., Pollak, T. A., McGuire, P., Fusar-Poli, P., … & David, A. S. (2020). Psychiatric and neuropsychiatric presentations associated with severe coronavirus infections: a systematic review and meta-analysis with comparison to the COVID-19 pandemic. The Lancet Psychiatry, 7(7), 611–627. doi:10.1016/S2215-0366(20)30203-0

13. de Sousa Moreira, J. L., Barbosa, S. M. B., Vieira, J. G., Chaves, N. C. B., Felix, E. B. G., Feitosa, P. W. G., … & Neto, M. L. R. (2021). The psychiatric and neuropsychiatric repercussions associated with severe infections of COVID-19 and other coronaviruses. Progress in Neuro-Psychopharmacology and Biological Psychiatry, 106, 110159. doi: 10.1016/j.pnpbp.2020.110159

14. Editorial COVID-19 and mental health. Lancet Psychiatry, 8 (2) (February 01, 2021). doi: 10.1016/S2215-0366(21)00005-5

15. Taquet, M., Luciano, S., Geddes, J. R., & Harrison, P. J. (2021). Bidirectional associations between COVID-19 and psychiatric disorder: retrospective cohort studies of 62 354 COVID-19 cases in the USA. The Lancet Psychiatry, 8(2), 130–140. doi: 10.1016/S2215-0366(20)30462-4

16. Ferrando, S. J., Klepacz, L., Lynch, S., Tavakkoli, M., Dornbush, R., Baharani, R., … & Bartell, A. (2020). COVID-19 psychosis: a potential new neuropsychiatric condition triggered by novel coronavirus infection and the inflammatory response?. Psychosomatics, 61(5), 551. doi: 10.1016/j.psym.2020.05.012

17. Caan, M. P., Lim, C. T., & Howard, M. (2020). A case of catatonia in a man with COVID-19. Psychosomatics, 61(5), 556. doi: 10.1016/j.psym.2020.05.021

18. Kopishinskaia, S., Cumming, P., Karpukhina, S., Velichko, I., Raskulova, G., Zheksembaeva, N., … & Smirnova, D. (2021). Association between COVID-19 and catatonia manifestation in two adolescents in Central Asia: incidental findings or cause for alarm?. Asian Journal of Psychiatry, 102761. doi: 10.1016/j.ajp.2021.102761

19. Mao, L., Jin, H., Wang, M., Hu, Y., Chen, S., He, Q., … & Hu, B. (2020). Neurologic manifestations of hospitalized patients with coronavirus disease 2019 in Wuhan, China. JAMA neurology, 77(6), 683–690. doi: 10.1001/jamaneurol.2020.1127

20. Tan, Y. K., Goh, C., Leow, A. S., Tambyah, P. A., Ang, A., Yap, E. S., … & Tan, B. Y. (2020). COVID-19 and ischemic stroke: a systematic review and meta-summary of the literature. Journal of thrombosis and thrombolysis, 50(3), 587–595. doi: 10.1007/s11239-020-02228-y

21. Rifino, N., Censori, B., Agazzi, E., Alimonti, D., Bonito, V., Camera, G., … & Sessa, M. (2021). Neurologic manifestations in 1760 COVID-19 patients admitted to papa Giovanni XXIII hospital, Bergamo, Italy. Journal of neurology, 268(7), 2331–2338. doi: 10.1007/s00415-020-10251-5

22. Diez□Quevedo, C., Iglesias□González, M., Giralt□López, M., Rangil, T., Sanagustin, D., Moreira, M., … & Cuevas□Esteban, J. (2021). Mental disorders, psychopharmacological treatments, and mortality in 2150 COVID□19 Spanish inpatients. Acta Psychiatrica Scandinavica, 143(6), 526–534. doi: 10.1111/acps.13304

23. Castilla, J., Guevara, M., Miqueleiz, A., Baigorria, F., Ibero-Esparza, C., Navascués, A., … & Ezpeleta, C. (2021). Risk Factors of Infection, Hospitalization and Death from SARS-CoV-2: A Population-Based Cohort Study. Journal of Clinical Medicine, 10(12), 2608. doi: 10.3390/jcm10122608

24. Chumakov, E., Petrova, N., & Smirnova, I. (2019). The evolution of views on mental disorders in patients with syphilis. Klinicheskaya Dermatologiya I Venerologiya, 18(1), 71. doi: 10.17116/klinderma2019180117125.

25. Pai, S., & Bader, G. D. (2018). Patient similarity networks for precision medicine. Journal of molecular biology, 430(18), 2924–2938. doi: 10.1016/j.jmb.2018.05.037

26. Huys, Q. J., Moutoussis, M., & Williams, J. (2011). Are computational models of any use to psychiatryã. Neural Networks, 24(6), 544–551. doi: 10.1016/j.neunet.2011.03.001

27. Friston, K. J., Redish, A. D., & Gordon, J. A. (2017). Computational nosology and precision psychiatry. Computational Psychiatry, 2–23. doi: 10.1162/cpsy_a_00001

28. Frässle, S., Aponte, E. A., Bollmann, S., Brodersen, K. H., Do, C. T., Harrison, O. K., … & Stephan, K. E. (2021). TAPAS: an open-source software package for Translational Neuromodeling and Computational Psychiatry. Frontiers in Psychiatry, 12, 857. doi: 10.3389/fpsyt.2021.680811

